# Cancer Variant Interpretation Group UK (CanVIG-UK): updates on an exemplar national subspecialty multidisciplinary network

**DOI:** 10.64898/2026.03.17.26348157

**Authors:** Alice Garrett, Sophie Allen, Charlie F Rowlands, Subin Choi, Miranda Durkie, George J Burghel, Rachel Robinson, Alison Callaway, Joanne Field, Bethan Frugtniet, Sheila Palmer-Smith, Jonathan Grant, Judith Pagan, Trudi McDevitt, Lowri Hughes, Elizabeth Johnston, Laura Yarram-Smith, Peter Logan, Laura Reed, Katie Snape, Helen Hanson, Terri McVeigh, Clare Turnbull, CanVIG-UK

## Abstract

Cancer Variant Interpretation Group UK was established in 2017 in response to the publication of the 2015 ACMG/AMP v3 guidance for the interpretation of sequence variants. Its initial purpose was to ensure consistency in the UK clinical-laboratory community implementation of ACMG/AMP v3 guidance for cancer susceptibility genes (CSGs). Still convening for monthly national meetings, the remit of CanVIG-UK now encompasses additional activities delivered under the following objectives:

Creation of a national multidisciplinary professional network and regular forum.Delivery of training and education.Establishment of a consensus approach to the fundamentals of variant interpretation in cancer susceptibility genes.Development and ratification of gene-specific frameworks for variant interpretation for cancer susceptibility genes.Development and maintenance of an online platform to facilitate information sharing and variant interpretation within the UK clinical-laboratory community.Facilitation of UK contribution to international variant interpretation endeavours.

A survey of CanVIG-UK members evaluating the impact of these activities conducted in November 2025 had 163 responses, including 113 clinical scientists/trainees and 27 Clinical Genetics consultants/trainees. The utility of the CanVIG-UK consensus recommendations for variant interpretation in cancer susceptibility genes was highly rated, with 89/145=61.4% of survey respondents reporting using the guidance at least weekly (≥4 times/month) and 124/128=96.9% rating it as extremely/very useful. The usage frequency and utility of the gene-specific guidance reported by survey respondents were similar to those reported for the main consensus specification. Both qualitative and quantitative survey responses clearly demonstrate the value of the CanVIG-UK activities to the clinical-diagnostic community.

**Key messages:**

- What is already known on this topic: Cancer Variant Interpretation Group UK (CanVIG-UK) is a national subspeciality multidisciplinary network first established in 2017. It brings together members of the UK clinical-laboratory community to improve accuracy and consistency in the interpretation of variants in cancer susceptibility genes (CSG)
- What this study adds: this article presents the results of a survey of CanVIG-UK members, demonstrating the impact of CanVIG-UK activities on their services, as well as a review of progress in the six updated objectives of CanVIG-UK
- How this study might affect research, practice or policy: this article presents current priorities and practices and potential future directions for variant interpretation in CSGs across the UK and Republic of Ireland

## Background

### The evolving landscape of variant interpretation for cancer susceptibility genes

With the advent of next generation sequencing and molecular technologies of increasingly higher throughput has come a dramatic increase in the volume of genetic testing offered for a variety of indications(1). This is likely to increase only further with the potential expansion of whole genome sequencing (WGS) and population-based analyses (2-5). Concurrent to this increase in generated genomic data is a need for more systematic, accurate and consistent approaches to the clinical interpretation of the variants output from bioinformatic pipelines.

The American College of Medical Genetics/Association for Molecular Pathology (ACMG/AMP) version 3 (v3) guidelines for the interpretation of sequence variants, published in 2015, significantly improved international consistency in variant interpretation by diagnostic labs(6). However, due to the wide scope of these guidelines, need for further specification and additional guidance for specific clinical settings became apparent. International variant curation expert panels (VCEPs) and working groups were tasked by ClinGen with providing gene-specific guidance that incorporates nuanced gene-specific information, including various VCEPs within the Hereditary Cancer Clinical Domain Working group domain focussing on cancer susceptibility genetics(7). Variant interpretation practice is constantly evolving as understanding and knowledge change, with the initiation of the development of new versions of guidance occurring often concurrently with the release of the previous version. The upcoming release of ACMG/AMP/College of American Pathologists (CAP)/ClinGen guidelines for the interpretation of sequence variants version 4 (v4) guidance will require substantial additional adaptions to current practice, requiring the restructuring of guidance to align with new frameworks.

### The value of subspecialty genomics networks

Different disease areas and their corresponding genes present different challenges, for example how phenotypic data should be used, taking into account penetrance and phenocopy rates. The interpretation of specialist clinical phenotype data, for example ophthalmological examination, ECG or neuroimaging findings, can require the involvement of clinical experts. The role of functional domains and understanding of mechanisms of disease can also vary gene by gene. The ACMG/AMP v3 guidance provides an overarching framework for variant interpretation, but just as different medical disciplines require their own specialist teams, genomic laboratories may assign different sub-teams to cover different disease areas e.g. cardiac genetics, neurogenetics. Achieving alignment in variant interpretation within each subspeciality area requires input both from clinical scientists with specialist expertise and clinicians with experience of relevant diseases.

### Cancer Variant Interpretation Group UK (CanVIG-UK)

In the UK NHS, regional molecular genetic laboratories with associated Clinical Genetics services are aligned into seven Genomic Laboratory Hubs (GLHs) covering the entirety of England (8). There are equivalent regional laboratory and clinical coverage across the other devolved nations of the UK. Families often span multiple regions and a consistent, equitable approach across the country is essential. The UK Association of Clinical Genomic Science (ACGS) has provided additional specifications of the ACMG/AMP v3 guidance for NHS implementation, primarily focussed on the rare disease setting(9). Due to the differences in the associated phenotypes and testing strategies, there were clear areas for which an approach specified for cancer susceptibility genes was also required. Furthermore, an array of cancer susceptibility genes were already routinely tested in NHS diagnostic laboratories for which interim national guidance was urgently required whilst awaiting outputs from the international VCEP groups. Cancer Variant Interpretation Group UK (CanVIG-UK) was established in 2017 with the specific primary aim of bringing the clinical-laboratory community together to implement the ACMG/AMP v3 guidance for cancer susceptibility genes, including reaching consensus on areas of subjectivity in the original guidance, as well as establishing a national sub-speciality multidisciplinary meeting(10).

Since its initiation, CanVIG-UK has served as a forum for professionals from different backgrounds to share expertise, enabling informed, multidisciplinary consensus on topical and complex areas of clinical variant interpretation practice for cancer susceptibility genes(10). CanVIG-UK membership-wide virtual meetings continue to take place monthly, with CanVIG-UK Steering and Advisory Group (CStAG) meetings also occurring monthly, focussing on the details of CanVIG-UK strategy, guidance and other resources.

In our previous publication in 2020, we set out the objectives of CanVIG-UK and presented survey data on CanVIG-UK membership and the perceived utility of the group functions for members at that time(10). Five years on, membership of the group has increased approximately four-fold and its remit has evolved, with its updated objectives shown in Box 1/Figure 1.

**Figure 1.**
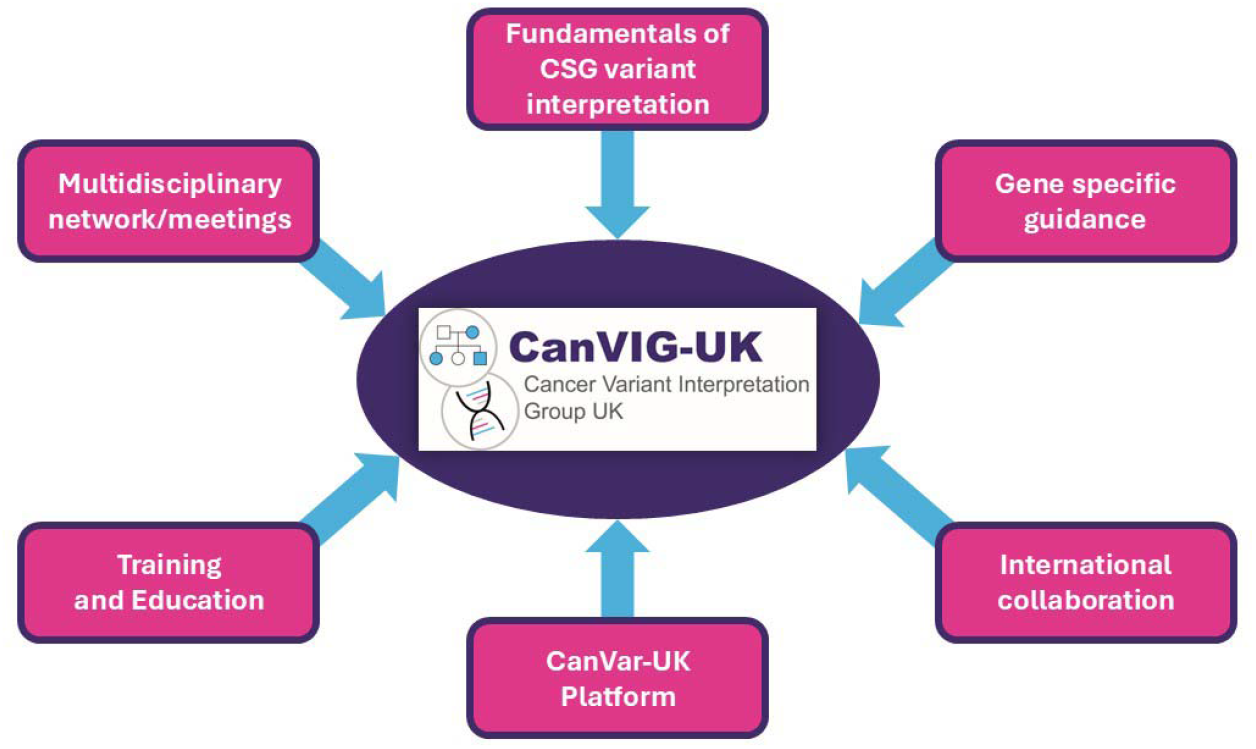
The six key objectives of CanVIG-UK in 2025/2026

#### Box 1

CanVIG-UK objectives 2025/2026

1. Creation of a national multidisciplinary professional network and regular forum.
2. Delivery of training and education.
3. Establishment of a consensus approach to the fundamentals of variant interpretation in cancer susceptibility genes.
4. Development and ratification of gene-specific frameworks for variant interpretation for cancer susceptibility genes.
5. Development and maintenance of an online platform to facilitate information sharing and variant interpretation within the UK clinical-laboratory community.
6. Facilitation of UK contribution to international variant interpretation endeavours.

In this article, we present the results of an updated and more comprehensive survey of members exploring how CanVIG-UK activities and resources influence their day-to-day clinical practice; we also discuss progress across the six CanVIG-UK objectives.

## Methods

The online survey was developed on the SurveyMonkey platform and was piloted on CStAG members before wider circulation to the CanVIG-UK membership. It comprised 20 questions relating to CanVIG-UK meetings and resources (see Supplementary table 1). Survey respondents were CanVIG-UK members who self-selected to participate after a verbal introduction to the survey in the November 2025 CanVIG-UK monthly meeting. A link to the survey was circulated both during the meeting and afterwards by email to all CanVIG-UK members. The survey remained open from 14^th^-28^th^ November 2025. Quantitative analyses were performed using R v4.4.1.

## Results

### Professional roles

163 CanVIG-UK members working in the UK and Republic of Ireland responded to the electronic survey invitation, of which 113/163=69.3% were self-reported qualified or trainee clinical scientists and 27/163=16.6% were consultant or trainee clinical geneticists (see Figure 2a). The roles of all survey respondents and their local genetics centres are in Supplementary tables 2 and 3. Approximately 2/3 survey respondents reported cancer genetics to be the sole or predominant aspect of their work (see Figure 2b).

**Figure 2.**
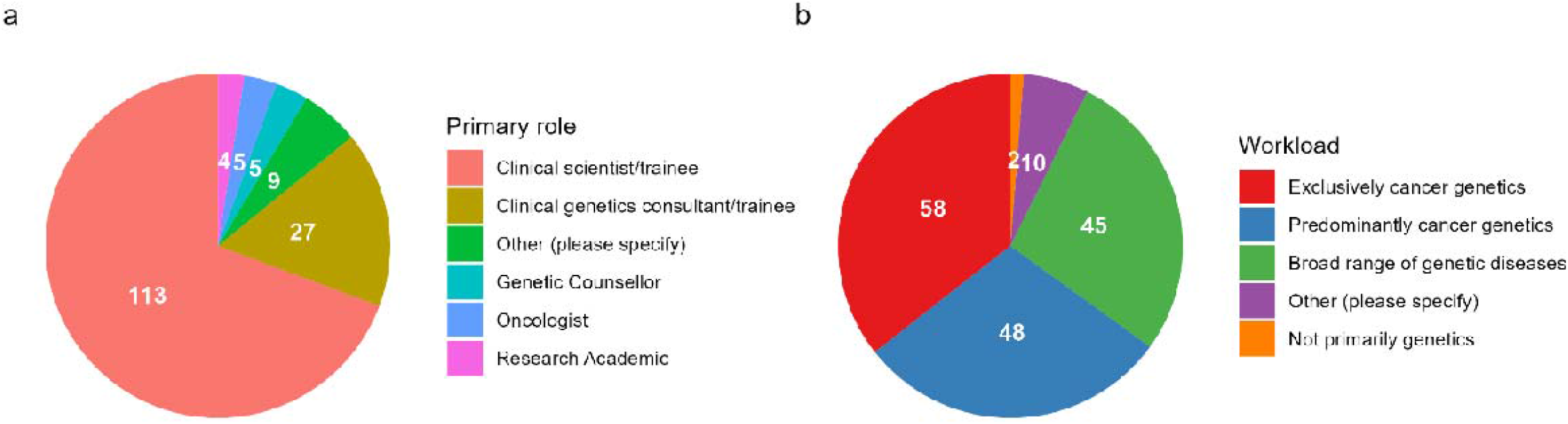
Pie-charts showing roles of survey respondents. a) Primary professional role, b) Nature of workload

### CanVIG-UK meetings

101/151=66.9% of survey respondents reported attending ≥5 of the eight CanVIG-UK meetings held annually (meetings being monthly with four omitted for vacations). The majority of meeting attendances were live rather than viewings of recordings (see Figure 3a and b). The reported motivations contributing to meeting attendance are summarised in Figure 3c, the highest-ranked motivations being ‘keeping up to date’ and ‘training/CPD’. The perceived relative utility by respondents of topic types covered in CanVIG-UK meetings is presented in Figure 3d, with “review of variant classification guidance” and “training demonstrations of new resources” being deemed of greatest utility. We also repeated analysis of survey responses restricted to clinical scientists/trainees (Supplementary Figure 1).

**Figure 3.**
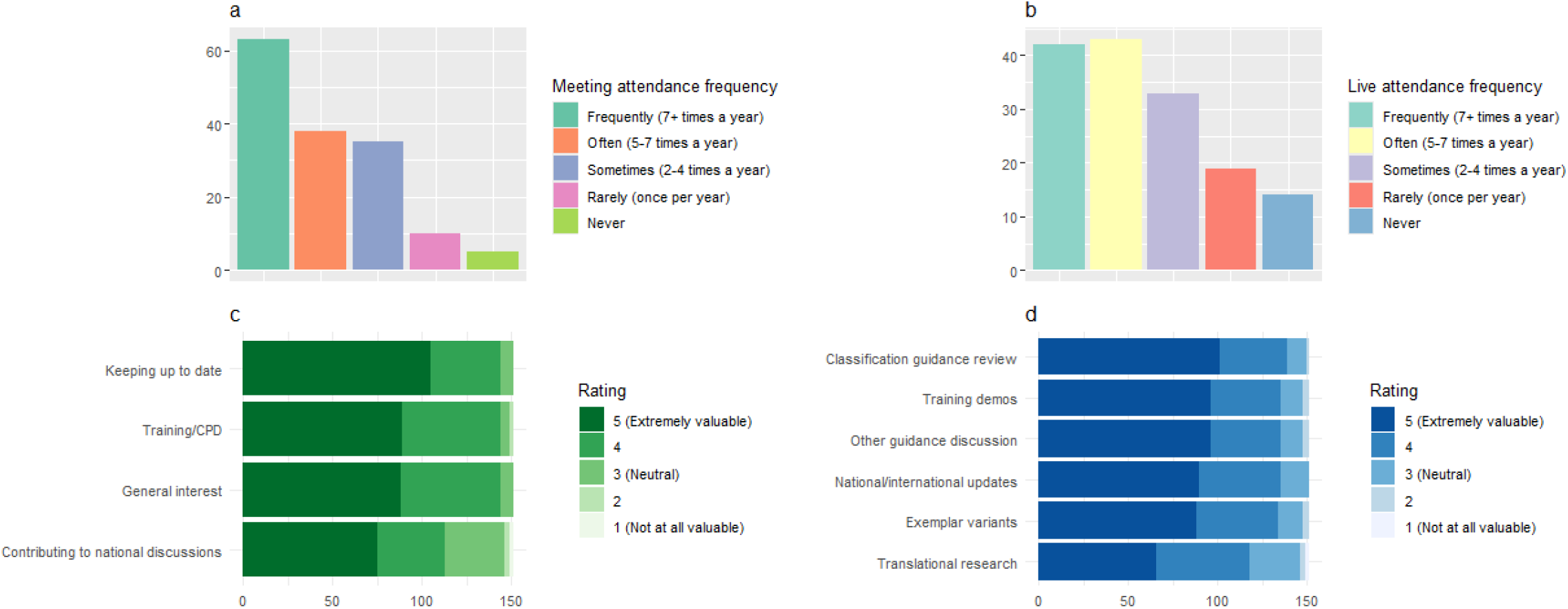
Bar charts showing counts of survey responses from all respondents on a) frequency of CanVIG-UK meeting attendance (either live or via meting recording), b) frequency of live CanVIG-UK meeting attendance, c) impact of different factors motivating attendance at CanVIG-UK meetings (each motivator rated 1-5), d) value of different topics discussed at CanVIG-UK meetings (each topic rated 1-5)

### CanVIG-UK resources and guidance

With regard to the main variant interpretation resources developed by CanVIG-UK, Figure 4a depicts for each resource the perceived utility and the frequencies with which these were used, with the consensus and gene-specific specifications scoring most highly (Figure 4a-b). With regard to specific challenging topics for which additional guidance was provided by CanVIG-UK, the perceived utility is presented in Figure 4c. The equivalent survey results on CanVIG-UK resources for responses from clinical scientists/trainees only are presented in Supplementary Figure 2.

**Figure 4.**
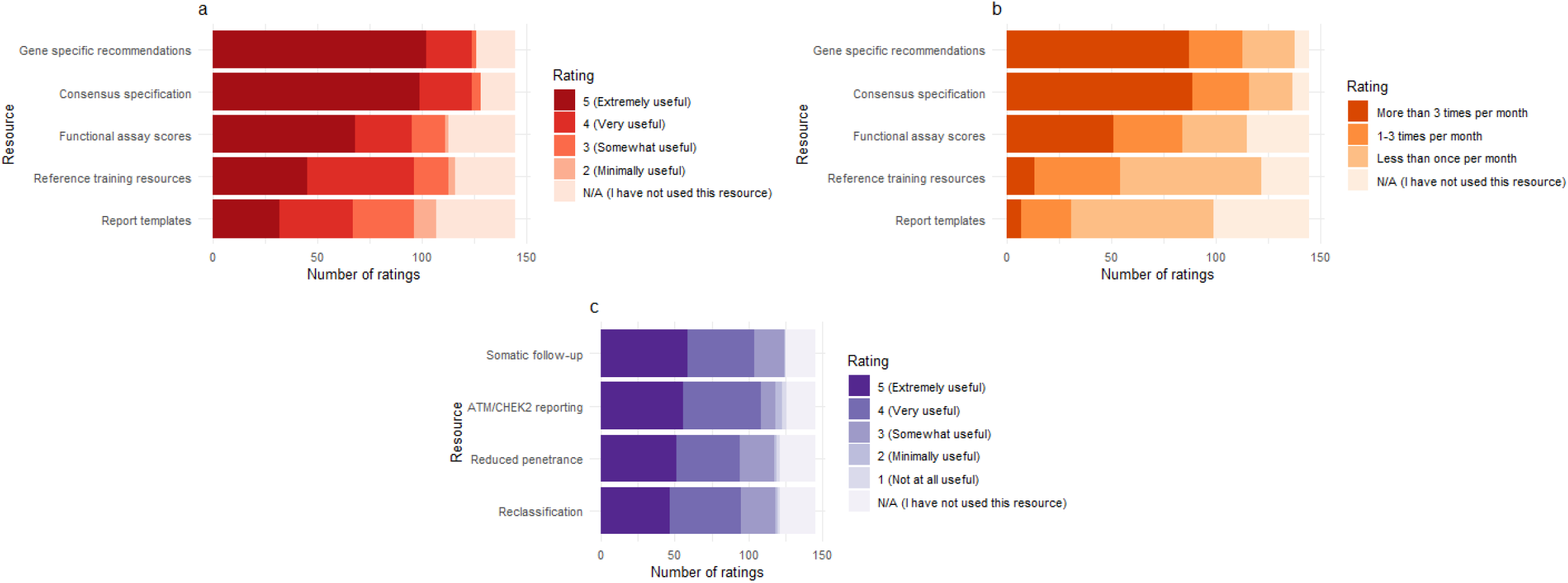
Bar charts showing counts of survey responses from all respondents on a) utility of core CanVIG-UK resources/guidance (each resource rated 1-5), b) frequency of usage of core CanVIG-UK resources/guidance meeting attendance (either live or via meting recording), c) utility of additional CanVIG-UK resources on specific challenging topics (each rated 1-5)

### CanVar-UK

When compared to Alamut, ClinVar, Franklin and Varsome, 91/137 = 66.4% of survey respondents reported CanVar-UK to be the resource they found most useful for assembling evidence for variant interpretation in cancer susceptibility genes, with a further 21.2% (29/137) placing it second. The median rating of the importance of the CanVar-UK platform for respondents’ work was 9/10. A comprehensive description of the CanVar-UK platform (the online cancer susceptibility gene variant interpretation platform developed and maintained by CanVIG-UK) with additional survey responses regarding its usage and utility are presented separately(11).

### Qualitative responses

41/163=25.2% of survey respondents provided additional free-text responses (Supplementary Tables 4 and 5). Notable amongst free-text responses were articulations of the substantial extent to which CanVIG-UK activities assist them in their professional role. Several responses from clinicians reflected that although they did not regularly attend CanVIG-UK meetings or use the CanVIG-UK resources themselves, they were aware of the important of the CanVIG-UK infrastructure is to the clinical scientists with whom they work, and thus commented affirmatively as to the indirect impact CanVIG-UK was having on their clinical service.

## Discussion: review of CanVIG-UK objectives

### Creation of a national multidisciplinary professional network and regular forum

About 2/3 of respondents in the present survey of CanVIG-UK members were clinical scientists. The current CanVIG-UK e-mail list comprises 466 individuals, of which 251 are clinical scientists in the UK and Republic of Ireland. The group continues to have monthly online meetings eight times per year (breaking for Christmas, Easter and summer holidays). The majority of survey respondents reported attending CanVIG-UK meetings ≥7 times a year (see Figure 2a). A high proportion of these reported attendances were live (see Figure 2b), indicating that this is most participants’ preferred option, but links to the meeting recordings are circulated after the meetings for those unavailable at the time of the meeting.

Meetings include internal and external speakers and cover a range of topics, responsive to the training needs of members as well as topical national and international issues. Most survey respondents rated “keeping up-to-date” as their main motivator for attending CanVIG-UK meetings, but “training/CPD” and “general interest” were also highly rated. Exemplar variants illustrative of meeting themes are circulated ahead of the meeting for pre-meeting classifications; the pre-meeting submissions are then collated and summarised for review during the live meeting. When establishing CanVIG-UK consensus approaches to complex issues, in-meeting polls and/or pre/post-meeting surveys are regularly used to capture opinions and experience from the wider CanVIG-UK membership. Agendas from recent CanVIG-UK meetings are presented in Supplementary Table 6 as exemplars.

Topics regularly covered at CanVIG-UK meetings uniformly received high utility ratings (see Figure 3d). These included reviews of variant classification guidance (including CanVIG-UK guidance and Variant Curation Expert Panel guidance), detailed classifications of exemplar variants, discussion of other national guidance (e.g. *ATM/CHEK2* reporting, reduced penetrance, reclassification), training demonstrations of relevant resources (e.g. COOL Bayes segregation analysis tool, quantitative assessment of functional assay), updates on translational research activities (e.g. PS4 likelihood ratio calculator, multiplexed assays of variant effects (MAVEs)), updates on national/international developments and working groups e.g. Somatic Variant Interpretation Group UK (SVIG), ClinGen functional working group, European Society for Medical Oncology. There were recurrent comments in the qualitative survey responses that equivalent infrastructure and regular meetings would be extremely useful for the interpretation of somatic variants (see Supplementary Table 5).

### Delivery of training and education

Training and education remain a priority and key role for CanVIG-UK, with high training utility ratings for the meetings themselves (see Figures 3c and 3d), as well as comments on the training utility in the qualitative survey responses (see Supplementary Table 4). The training needs of CanVIG-UK members are the drivers of meeting content, and requests from survey respondents on future CanVIG-UK meeting topics are shown in Supplementary Table 5. Additionally, CStAG and CanVIG-UK members regularly contribute in a breadth of settings to educational delivery regarding variant interpretation on cancer susceptibility genes, including courses and workshops through the national genomic training academy (GTAC), massive open online courses (MOOCs), external quality assessment (EQA) schemes and external courses (12-15).

### Establishment of a consensus approach to the fundamentals of variant interpretation in cancer susceptibility genes

Twelve iterations of the CanVIG-UK consensus specification for the interpretation of variants in cancer susceptibility genes have been published on the CanVIG-UK website (https://www.cangene-canvaruk.org/canvig-uk) since 2021; thus is reflective of the rate of emergence of new evidence, published international guidance and consensus methodologies. 89/145=61.4% of survey respondents reported using the guidance at least weekly (≥4 times/month) and 124/128=96.9% of those using it rated it as extremely/very useful. There were 12,365 page views of the consensus specification page on the CanVIG-UK website in the past year (25/2/25-24/2/26, https://www.cangene-canvaruk.org/canvig-uk-guidance).

In addition, CanVIG-UK has developed consensus frameworks for the approaches to specific challenging clinical scenarios, including (i) variant reclassification, (ii) classification of variants of reduced penetrance, (iii) germline follow-up of variants found on tumour (somatic) testing and (iv) reporting of variants in moderate risk genes such as *ATM* and *CHEK2* (16-20). These frameworks were developed iteratively, informed by examples from the CanVIG-UK community, surveys of CanVIG-UK members and multiple discussions at CanVIG-UK meetings. Frameworks are typically developed jointly with the UK Cancer Genetics Group (UKCGG), the national multidisciplinary organisation focused on improving the clinical care of individuals with a hereditary predisposition to cancer, ensuring applicability to the clinical cancer genetics community. The majority of survey respondents rated the utility of each of these additional guidance documents as 4/5 (very useful) or 5/5 (extremely useful) (see Figure 3c). There were 1,882 page views of the resources page on the CanVIG-UK website where these frameworks are displayed in the past year (25/2/25-24/2/26, https://www.cangene-canvaruk.org/canvig-uk-variant-resources).

Also presented on the CanVIG-UK website are report templates, recommendations for the application of evidence from functional assays and resources from previous CanVIG-UK sessions. The frequency with which survey respondents reported using these resources varied, but high proportions of those had used the resources reporting their utility to be very useful (4/5) or extremely useful (5/5) (see Figure 3a). An index of the CanVIG-UK resources shown in figures 3a-c are shown in supplementary table 7.

CanVIG-UK collaborates closely with the UK Somatic Variant Interpretation Group (SVIG-UK), who have recently published a UK consensus approach for interpretation of somatic variants identified through genomic testing in patients with solid tumours and haematological malignancies(21, 22). CanVIG-UK regularly hosts joint meetings with SVIG-UK colleagues to discuss issues relevant to the entire community.

### Development and ratification of gene-specific frameworks for variant interpretation for cancer susceptibility genes

In addition to the general consensus specification relevant to all cancer susceptibility genes, the CanVIG-UK website now displays 14 sets of gene-specific recommendations covering 20 genes to be used alongside the general consensus specification. These typically have been developed within CanVIG-UK to serve as interim guidance whilst awaiting development of VCEP guidance. On release of the corresponding VCEP recommendations, detailed review and harmonisation is performed, augmenting with additional specification as required based on feedback from CanVIG-UK members. Points of uncertainty are clarified through correspondence with the VCEP coordinators. 87/145=60.0% of survey respondents reported using the gene-specific guidance at least weekly (≥4 times/month) and 124/126=98.4% of those using it rated it as extremely/very useful (see Figures 3a and 3b). There were 16,335 page views of the gene-specific recommendations page on the CanVIG-UK website in the past year (25/2/25-24/2/26, https://www.cangene-canvaruk.org/gene-specific-recommendations).

CanVIG-UK has also published novel quantitative analyses and methodologies relating to evidence application of case-control, in silico, functional and phenotypic-specificity data; these have been incorporated into the CanVIG-UK general consensus and gene-specific variant interpretation recommendations(23-27).

### Development and maintenance of an online platform to facilitate information sharing and variant interpretation within the UK clinical-laboratory community (CanVar-UK)

A full description of the CanVar-UK platform and its usage and perceptions regarding its utility are described elsewhere(11). The data displayed and functions provided by CanVar-UK have been designed in specific response to input from CanVIG-UK members and to service the requirements of the NHS clinical scientist community. Previously, variant-level discussions and information-sharing between CanVIG-UK members was undertaken via email; this has been superseded by a diagnostic discussion forum through which all comments are captured in CanVar-UK against the relevant variant. The forum also provides the infrastructure for urgent reclassification alerts to be disseminated to the community. The diagnostic user forum and other CanVar-UK functions are widely used by CanVIG-UK members and high utility ratings and positive qualitative comments were given in the survey (see Supplementary tables 3 and 4)(11) Initially limited to NHS employees, membership of the forum has now been expanded to international users from clinical diagnostic laboratories.

### Facilitation of UK contribution to international variant interpretation endeavours

Multiple members of CStAG contribute to international groups working on variant interpretation in cancer susceptibility genes, providing channels for communication with the CanVIG-UK community. These include the Evidence-based Network for the Interpretation of Germline Mutant Alleles (ENIGMA) *BRCA1/2* VCEP and steering committee, the Hereditary Breast, Ovarian and Pancreatic cancer VCEP, the Endocrine Tumor Predisposition VCEP, Neurofibromatoses and Schwannomatosis VCEP, the Ocular Oncology VCEP, the Myeloid Malignancy VCEP, the ClinGen functional working group and the Atlas of Variant Effect Alliance. CStAG members also regularly give invited talks at international conferences and meetings.

CanVIG-UK meetings are an opportunity to share and discuss international updates with the UK clinical-diagnostic laboratory community and 135/151=89.4% of survey respondents rated national and international updates at CanVIG-UK meetings as very or extremely useful. All CanVIG-UK resources are freely available to the international community through the CanVIG-UK website and during the past year there were 23,147 site sessions (website visits) by 2,041 unique visitors from 80 countries across all continents (25/2/25-24/2/26). Of inquiries to the CanVIG-UK e-mail address regarding CanVIG-UK resources, >10% are from users based outside the UK.

## Conclusions

The expanding role of CanVIG-UK in facilitating coordinated and consistent approaches to variant interpretation in cancer susceptibility genes across the UK and ROI, as demonstrated by the activities centred around the five outlined objectives, is supported by high engagement from the clinical-diagnostic community. Far from variant interpretation being a “solved problem”, the increased complexity of international variant interpretation guidance, data and tools have resulted in an even greater need for community collaboration to improve consistency. The affirmative responses from the surveyed CanVIG-UK members regarding the utility of CanVIG-UK activities, demonstrated particularly by the qualitative free-text responses, show how critical CanVIG-UK endeavours have been to its members.

The most imminent challenge facing the clinical-diagnostic community is likely to be the practical implementation of ACMG/AMP/CAP/ClinGen guidelines for the interpretation of sequence variants version 4 (v4) guidance. Nevertheless, the role of CanVIG-UK is likely impactful well beyond this, as resources, knowledge and technology will continue to evolve. CanVIG-UK is currently the only subspeciality network of its kind in the UK and ROI, but groups with equivalent infrastructure would likely offer equivalent value in other subspecialty disease areas, such as cardiac genetics and neurogenetics.

## Data Availability

All data produced in the present work are contained in the manuscript

## Acknowledgements

A full list of CanVIG-UK consortium members and their affiliations appears in the Supplemental Material. SA and CR are funded by CG MAVE (CRUK Programme Award [EDDPGM-Nov22/100004]). AG receives funding from NHS England. HH is supported by the National Institute for Health and Care Research Exeter Biomedical Research Centre (NIHR203320). CanVIG-UK activities 2019-2025 were funded by the CRUK Catalyst Award CanGene-CanVar (C61296/A27223) 2019-2024

