## Supplementary figures for "Cancer Variant Interpretation Group UK (CanVIG-UK): updates on an exemplar national subspecialty multidisciplinary network"

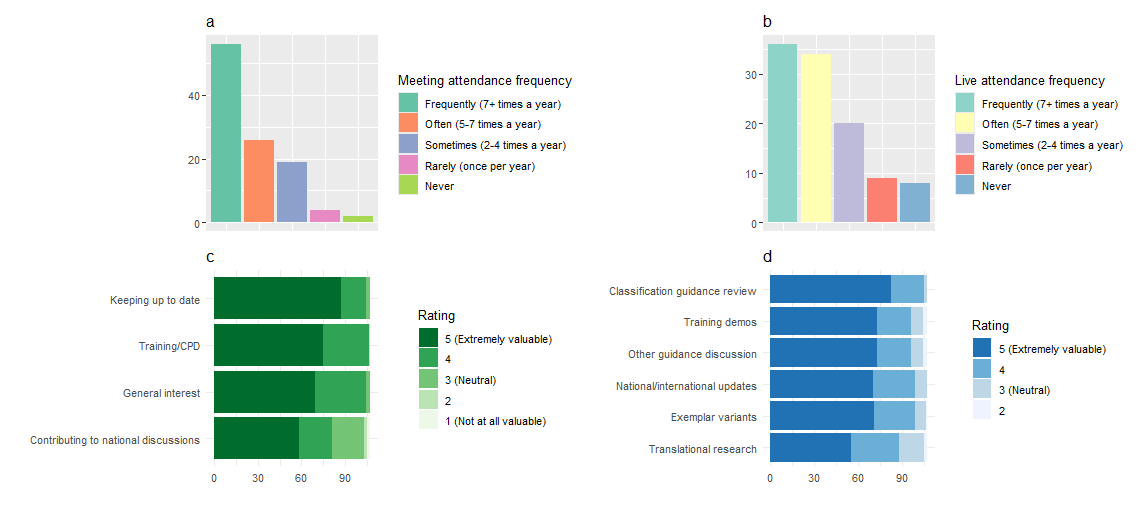

**Supplementary Figure 1:** Bar charts showing survey responses from 113 participants with a primary role of ‘Qualified Clinical Scientist’ or ‘Clinical Scientist Trainee’. Questions pertain to a) frequency of CanVIG-UK meeting attendance (either live or via meting recording), b) frequency of live CanVIG-UK meeting attendance, c) effect of different factors on motivation to attend CanVIG-UK meetings (each motivator rated 1-5), d) value of different topics discussed at CanVIG-UK meetings (each topic rated 1-5)

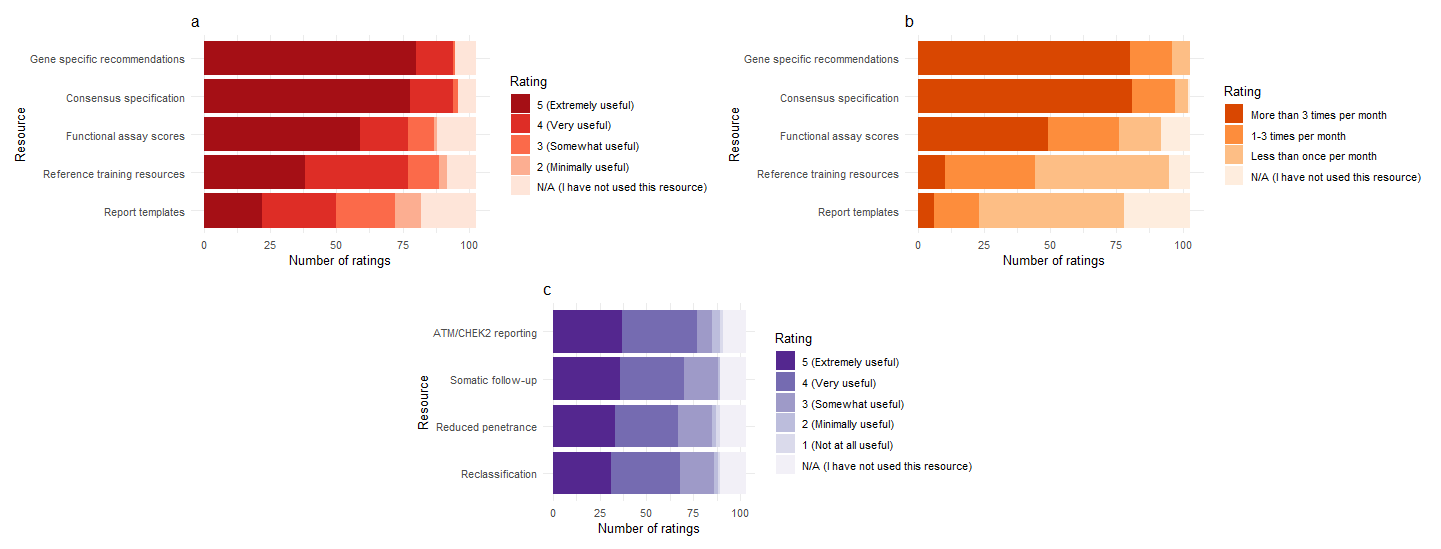

**Supplementary Figure 2:** Bar charts showing survey responses from 113 participants with a primary role of ‘Qualified Clinical Scientist’ or ‘Clinical Scientist Trainee’. Questions pertain to a) utility of core CanVIG-UK resources/guidance (each resource rated 1-5), b) frequency of usage of core CanVIG-UK resources/guidance meeting attendance (either live or via meting recording), c) utility of additional CanVIG-UK guidance on specific challenging topics (each rated 1-5)
